# Preliminary Report of Academic CAR-T (ISIKOK-19) Cell Clinical Trial in Turkey: Characterization of Product and Outcomes of Clinical Application

**DOI:** 10.1101/2021.09.23.21263731

**Authors:** Ebru Erdogan, Koray Yalcin, Cansu Hemsinlioglu, Aslihan Sezgin, Utku Seyis, Derya Dilek Kancagi, Cihan Tastan, Bulut Yurtsever, Raife Dilek Turan, Didem Cakirsoy, Selen Abanuz, Gozde Sir Karakus, Muhammer Elek, Huseyin Saffet Bekoz, Ali İhsan Gemici, Deniz Sargin, Mutlu Arat, Burhan Ferhanoglu, Ebru Pekguc, Serdar Ornek, Deram Buyuktas, Nur Birgen, Siret Ratip, Ercument Ovali

**Author notes:** These authors contributed equally to this work (Ebru Erdogan, Koray Yalcin).

## Abstract

**Objective:** Chimeric antigen receptor T (CAR-T) cell therapies already made an impact on the treatment of B cell malignancies. Although CAR-T cell therapies are promising, there are concerns with commercial products regarding their affordability and sustainability. In this preliminary study, results of the first productional and clinical data of academic CAR-T cell (ISIKOK-19) from Turkey are presented.

**Materials and Methods:** A pilot clinical trial (NCT04206943) designed to assess the safety and feasibility of ISIKOK-19 T-cell therapy in patients with relapsed and refractory CD19+ tumors was conducted and participating patients received ISIKOK-19 infusions between October 2019 and July 2021. Production data of the first 8 patients and the clinical outcome of 7 patients who received ISIKOK-19 cell infusion is presented in this study.

**Results:** Nine patients were enrolled for the trial (ALL n=5 and NHL n=4) but only 7 patients could receive the treatment. Two out of three ALL patients and three out of four NHL patients had complete/partial response (ORR 72%). Four patients (57%) had CAR-T-related toxicities (CRS, CRES, and pancytopenia). Two patients were unresponsive and had progressive disease following CAR-T therapy. Two patients with partial response had progressive disease during follow-up.

**Conclusion:** Production efficacy and fulfilling the criteria of quality control were satisfactory for academic production. Response rates and toxicity profiles are acceptable for this heavily pretreated/refractory patient group. ISIKOK-19 cells appear to be a safe, economical, and efficient treatment option for CD19 positive tumors. The findings of this study need to be supported by the currently ongoing clinical trial of ISIKOK-19.

## Introduction

Cellular therapies are promising to have a wider application in the treatment of malignancies in the near future. Especially, chimeric antigen receptor T (CAR-T) cell therapies already made an impact on the treatment of B cell malignancies. Based on the impressive clinical outcomes, there is an increasing consensus that CAR-T cells have the potential to deliver powerful antiLJtumor effects. These efforts culminated in the approval of CAR-T cells by the medical authorities and commercial products have been used worldwide.^1, 2^

A typical CAR structure consists of an extracellular domain for tumor antigen recognition, linked to one or more intracellular signaling domains that mediate T□cell activation. The antigen□binding single-chain variable fragment (scFv) portion of a CAR has the variable heavy (V_H_) and variable light (V_L_) chains of an antibody which are fused by a peptide spacer. This extracellular structure is joined to an intracellular signaling molecule comprised of the TCR CD3ζ signaling chain, and optional tandem co□stimulation domains such as the 4□1BB or CD28 signaling modules. CAR-T cell□mediated targeting of tumors is neither restricted nor dependent on antigen processing and presentation due to scFvs which possess the capacity to recognize intact cell surface proteins. Regarding this aspect, CAR-T cells are therefore unaffected by tumor escape mechanisms related to MHC loss.

The academic CAR-T product in this study, ISIKOK-19 cells, encode the anti-CD19 CAR construct with scFv of an anti-CD19 monoclonal antibody (FMC63) conjugated with the CD8 hinge region, CD28 transmembrane (TM), and co-stimulatory domain, and the CD3ζ pro-activator signaling domain along with a truncated form of epidermal growth factor receptor (EGFRt) cell surface protein as a co-expression marker and a safety switch mechanism.

Several factors related to production and clinical application can influence CAR-T cell treatment’s overall outcome. In the production phase, CAR-T cell quality is depended on the number and viability of T-cells, CD4:CD8 ratio, T-cell subsets, the extent of expression of CAR on the cell surface, cytokine production, presence of bacterial endotoxins, risk of insertional oncogenesis, presence of residual magnetic beads, and sterility. Also, there are clinical factors that include immune-dependent cancer antigen selection, preferably cancer-specific antigen; CAR-T persistence in the patient; and associated toxicity profile with cytokine release syndrome (CRS) and CAR-T related encephalopathy syndrome (CRES).

Commercial availability of the CAR-T products provides large real-world experience in treating relapsed or refractory B-cell malignancies, shed light on anti-CD19 CAR-T cell products’ feasibility and adverse effects such as CRS, CRES, and cytopenia. There are multiple ongoing clinical CAR-T cell therapy-based trials which include studies comparing CAR-T cell therapy to hematopoietic stem cell transplantation (HSCT) or investigating their use at earlier stages of the disease and novel combinations.^3-5^

Although CAR-T cell therapies are promising, there are concerns with commercial products regarding their affordability and sustainability in many countries. These concerns raised the need for more economical academic and local production. Hence, several countries are exploring alternative T cell production models which have a comparable clinical outcome to commercial products.^6^ In this preliminary study, results of the first productional and clinical data of academic CAR-T cell product ISIKOK-19 from Turkey are presented.

## Methods

A pilot clinical trial (NCT04206943) designed to assess the safety and feasibility of ISIKOK-19 T-cell therapy in patients with relapsed and refractory CD19+ tumors was conducted at Acıbadem Altunizade Hospital. Production and treatment protocol was approved by the respective institutional review board (56733164/203). All the patients provided written informed consent. Participating patients received ISIKOK-19 infusions between October 2019 and July 2021. Production data of the first 8 patients and the clinical outcome of 7 patients who received ISIKOK-19 cell infusion is presented in this study.

### ISIKOK-19 Cell Production

Patients were subjected to leukapheresis at the Apheresis Unit of Acıbadem Altunizade Hospital. Apheresis procedures were performed using Amicus device (Fresenius Kabi, Lake Zurich, IL). A minimum of 1×10^8^ total nucleated cells diluted in 50 ml of plasma was collected.

Following leukapheresis, CD3+ T-cells were isolated using anti-CD3 magnetic beads (Miltenyi) following isolation of PBMCs by overlaying blood on Ficoll-Paque PLUS (GE Healthcare). T-cell activation was performed with human anti-CD3/anti-CD28 Dynabeads (Thermo Fisher Scientific). The plasmids which composed the lentiviral vector encoding CD19-specific CAR (anti-CD19 scFv h(CD28-CD3ζ)-EGFRt), were designed and synthesized by Creative Biolabs and lentivirus was synthesized in Acıbadem Labcell Laboratory as described previously.^7^ Culture and expansion processes of transduced T-cells were within 11-14 days in complete T cell medium [50 U/mL interleukin (IL)-2, 10 ng/mL IL-7, 10 ng/mL IL-15, 3% human AB serum, and 1% pen/strep, TexMACS Medium]. The CAR expression level was determined by Miltenyi MACSQuant flow cytometry analysis using anti-EGFR-A488 antibody on CD3 cells (R&D Systems). Finally, CAR-T cell production was performed in GMP units of Acıbadem Labcell Laboratory.

### Quality Control Tests

The purity of the CAR+ cells was analyzed by estimating the percentage of CAR+ cells and CAR expression on cells was assessed using EGFRt+ CD3 specific antibodies as described before.^7^ And also Subset analysis of T cells was done and exemplified by the CD3 (APC-H7, BD 560176), CD4 (Pacific Blur, Biolegend 300521), CD8 (APC, BD 555369), CD45RA (PE-CY7, BD 337186), CD45RO (FITC, BD 555492), and CD62L (PE, BD 341012) expressions in samples from different patients. Subsets of T cells were defined as described before.^8^

Potency was assessed by two different methods; (1) levels of proinflammatory cytokines such as IFNg were observed when ISIKOK-19 cells were co-cultured with RAJI, in comparison with ISIKOK-19 cells alone (2) target cell lysis capacity was measured by flow cytometer detecting Raji cell lysis capacity of CAR+ cells. The cytotoxic potential was analyzed in vitro for each product before infusion. A co-culture of the final product with RAJI cell line was initiated at different E:T ratios. The percentage of alive-CD19+ cells was measured by flow cytometry after 4 hours. As a control, the cytotoxic activity of non-transduced CD4+ CD8+ cells from the same patient was also measured. Products were considered valid when both the RAJI cell death percentage at ratio 1:1 and CD25+ regarding CAR-T activation was greater than 80%.

Replicative competent lentivirus (RCL) safety test was performed with CAR-T cell supernatants, which were cultured weekly for 3 passages, and then HIV RNA PCR was used for detecting lentivirus replication.

Other quality control tests of the virus and CAR-T cells including live cell number, sterility (aerobic, anaerobic, fungal, and mycoplasma analyses), endotoxin level, transgene CAR copy number/cell analysis, impurity assay, stability assays, and tumorigenicity tests (including cytogenetic analysis and relative telomerase activity (RTA) (TeloTAGGG Telomerase PCR ELISA-Roche)) were performed in Acıbadem Labmed Laboratory by accredited methods and details of quality control tests as defined previously.^7^

### CAR-T Cell and CD19 Expression Monitoring

CAR-T cells were detected and followed by HIV RNA copy number/μDNA as described previously^7^ and the percentage of CD19 positive cells in peripheral blood were estimated by flow cytometer using anti-human CD19 (PE/Cy7, Biolegend 302216). Positive CD19 expression in the tissues was also detected histochemically by using CD19 monoclonal antibody.

### CAR-T cell Treatment

Following leukapheresis, three patients received interim therapy (bridge to CAR-T) at the discretion of the treating physician. All the patients received chemotherapy aimed at T lymphocyte depletion. The clinical treatment plan consisted of a course of lymphodepletion chemotherapy, followed 1 day later by infusion of anti-CD19 CAR-T cells. Lymphodepletion chemotherapy consisted of cyclophosphamide at a total dose of 300 mg/m^2^, followed by three daily doses of fludarabine 30 mg/m^2^. Chemotherapy was administered before anti-CD19 CAR-T cells to deplete endogenous lymphocytes which can inhibit the anti-malignancy activity of adoptively transferred T cells.^9^ Targeted dose of ISIKOK-19 cells was defined as 4-8 ×10^6^ CAR-positive T cells/kg. To avoid severe cytokine release syndrome split dose administration was preferred. The total cell amount was divided into three doses and was infused as 20% on day 0, 30% on day 2, and 50% on day 7.

Response evaluation was done according to FDG PET/CT analysis for NHL and according to bone marrow aspiration and FDG PET/CT (for extramedullary involvement) analysis for ALL. Toxicity analysis including CRS or CRES was evaluated according to Lee criteria.^10^

## Results

### CAR-T Cell Product Characterization

Nine apheresis products were obtained from 9 patients registered in the clinical trial. For one patient, ISIKOK-19 cell infusion could not be done due to production failure. Another patient died before cell production was completed. Characteristics of ISIKOK-19 for 8 patients, excluding the patient with production failure, are presented.

Apheresis products were subjected to CD3+ magnetic selection using the CliniMACS system. For all cases, the minimum required number of T-cells (149 × 10^6^) was obtained. Cells were expanded for an average of 10.5 days (range 9–12). The average total cell number obtained in the final product was 1436×10^6^ (range 164 to 3690×10^6^).

The final product was characterized in terms of cell viability, percentage of CD3+ cells, percentage of CAR+ cells, and targeted cell number. All products met acceptance criteria for cell viability, percentage of CD3+ cells, and percentage of CAR+ (>80% for cell viability, >95% for CD3 cells, and >20% for CAR+). The lowest value detected for cell viability was 88% for CD3+ cells. (Table-1)

**Table-1.** ISIKOK-19 product characterization

| Patient | Total cell Number (x10<sup>6</sup>) | CD3% | CAR% | Cell Viability | Sterility/ Mycoplasma | Impurity | Stability | Cytogenetic Analysis | RTA | Endotoxin | Transgene Copies/cell | RCL | POTENCY |
| --- | --- | --- | --- | --- | --- | --- | --- | --- | --- | --- | --- | --- | --- |
| P1 | 164 | 98.3 | 87 | 91 | Sterile | Passed | Passed | Normal | <20 | <0.5 EU/ml | 5 | Absent | >%80 |
| P2 | 3040 | 97.6 | 23.1 | 88 | Sterile | Passed | Passed | Normal | <20 | <0.5 EU/ml | 6 | Absent | >%80 |
| P3 | 628 | 98.8 | 32.5 | 98 | Sterile | Passed | Passed | Normal | <20 | <0.5 EU/ml | 4 | Absent | >%80 |
| P4 | 1164 | 99.1 | 29.4 | 94 | Sterile | Passed | Passed | Normal | <20 | <0.5 EU/ml | 2 | Absent | >%80 |
| P5 | 1148 | 96.7 | 28.8 | 92 | Sterile | Passed | Passed | Normal | <20 | <0.5 EU/ml | 2 | Absent | >%80 |
| P6 | 3690 | 99.9 | 21 | 96 | Sterile | Passed | Passed | Normal | <20 | <0.5 EU/ml | 8 | Absent | >%80 |
| P7 | 213 | 99.2 | 35.8 | 90 | Sterile | Passed | Passed | Normal | <20 | <0.5 EU/ml | 9 | Absent | >%80 |
| P8 | 1442 | 99.3 | 36 | 97 | Sterile | Passed | Passed | Normal | <20 | <0.5 EU/ml | 7 | Absent | >%80 |

Using EGFRt-specific antibody detection, the percentage of CAR+ (ISIKOK-19) cells in the patients’ products was assessed. All products met the specification of >20% ISIKOK-19 cells. Average percentage of CAR+ was 36.7% (range 21-87%). CAR19 transduction was also assessed in terms of DNA copies/cell. CAR19 was detected in all products, within a range of 2-9 copies/cell (all below the limit considered safe of <10 copies/cell). In terms of cytotoxicity, all products obtained met the specifications indicating that they had the capacity of eliminating CD19+ cells with the average percentage of 87% (range 83-92%).

Product composition was analyzed in terms of the CD4/CD8 ratio. As observed in the previous reports^6, 11^ CD4/CD8 ratio was inverted (CD4/CD8 ratio < 1) in all patients who were candidates for CAR-T cell therapy. Mean CD4/CD8 ratio was 0.39 (range 0.17-0.85) in apheresis products. This ratio was significantly altered after CD3+ cell selection for all patients, the mean CD4/CD8 ratio increased from 0,39 to 0,50. A significant increase was detected in the proportion of CD4+ cells after transduction and expansion, the mean ratio of EGFR positive CD4/CD8 ratio of the final product was detected as 1.75 (range 0.25-5.05). T cell subsets were identified regarding CD62L positivity to detect the total ratio of T-Naive (T_N_), T-Stem Cell Central Memory (T_SCM_) and T-Central Memory (T_CM_) mean 39.3% (range 11.7-71.8). Also, memory T cell dominancy was defined by CD45RO positivity mean 87.4% (range 70-99.1). (Table-2)

**Table-2.** CD4/CD8 Ratio of ISIKOK-19 in Different Production Steps and Subsets of T Cells

| Patient | CD4/CD8 Ratio |  |  | Subsets of T Cells in the Final Product |  |
| --- | --- | --- | --- | --- | --- |
|  | After Apheresis | After Selection | Final Product | CD62L +<br>(T<sub>CM</sub>, T<sub>SCM</sub>, T<sub>N</sub>) | CD45RO+<br>(Memory T Cell) |
| P1 | 0.38 | 0.98 | 5.05 | 71.8% | 97.8% |
| P2 | 0.17 | 0.52 | 0.61 | 42.3% | 99% |
| P3 | 0.49 | 0.35 | 0.25 | 11.7% | 76.4% |
| P4 | 0.45 | 0.24 | 2.13 | 50.9% | 93.4% |
| P5 | 0.18 | 0.48 | 0.84 | 41.4% | 76.4% |
| P6 | 0.85 | 0.52 | 2.61 | 15.8% | 99.1% |
| P7 | 0.25 | 0.47 | 0.79 | 41.5% | 70% |
| P8 | 0.42 | 0.57 | 0.81 | 43.7% | 82% |

### In Vivo Expansion and Persistence of ISIKOK-19

ISIKOK-19 sequences remained detectable, and quantitative polymerase-chain-reaction (PCR) showed very high levels of proliferation of ISIKOK-19 cells. All patients had peak levels greater than 3000 copies per microgram of genomic DNA, and five patients had peak levels greater than 30,000 copies per microgram of genomic DNA. One patient (Patient 3) had a peak value of only 3969 copies on the 14th day, quickly lost ISIKOK-19 cells subsequently, and had progressive disease. (Figure-1)

**Figure-1.**
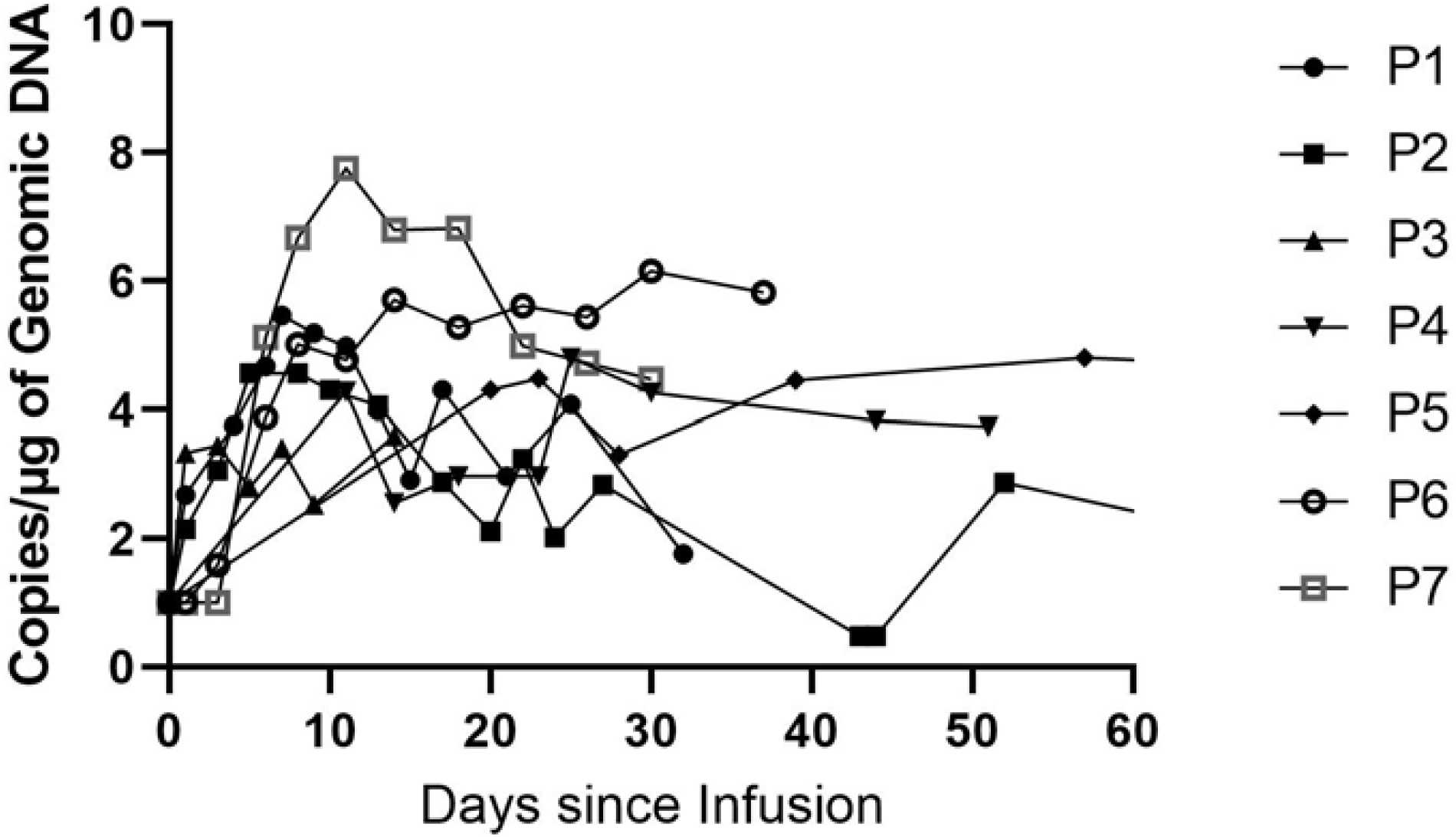
Measurements of ISIKOK-19 cells in peripheral blood as assessed by means of quantitative real-time polymerase-chain-reaction (PCR) assay

In patients with objective response (complete or partial), B-cell aplasia (absence of CD19-positive cells in peripheral blood) continued for a long duration of time, and CD19-positive cells were not detected in the biopsies of responsive lymphoma patients after CAR-T cell therapy at the time of progression. Also, CAR-T copy numbers lasted during follow-up in all responsive patients even after progression indicating progressive disease without CD19 positive cells in three patients. (Table-3)

**Table-3.** ISIKOK-19 Proliferation and B Cell Aplasia

|  | Peak Copies/ug DNA | Day of Peak Copies | The Last CAR-T Copy Number (TimePoint) | The Last Time Point of B Cell Aplasia | CD19 Expression In the Biopsy After CAR-T |
| --- | --- | --- | --- | --- | --- |
| P1 | 289934 | 7 | 14372<br>(10 months/ existing during follow-up) | 10 months<br>(during follow-up) | CD19 negative |
| P2 | 36792 | 8 | 4469<br>(12 months/ disappear after HSCT) | 4 months<br>(until HSCT) | NA |
| P3 | 3969 | 14 | Zero<br>(16th day of infusion) | No B cell aplasia | NA |
| P4 | 63266 | 25 | 5456<br>(2 months/ disappear after salvage therapy) | 58 days<br>(until salvage therapy) | NA |
| P5 | 281757 | 128 | 17550<br>(7 months/ existing during follow-up) | 7 months<br>(during follow-up) | CD19 negative |
| P6 | 1427090 | 30 | 659356<br>(2 months/ existing during follow-up) | 2 months<br>(during follow-up) | NA |
| P7 | 56678000 | 11 | 29444<br>(2 months/ existing during follow-up) | 2 months<br>(during follow-up) | NA |

### Clinical Outcomes

Three patients with DLBCL, three patients with ALL, and one with double-hit lymphoma (DHL) were treated. (Table-4) One patient had DLBCL transformed from CLL and the patient with DHL transformed from follicular lymphoma. All four lymphoma patients had the chemotherapy-refractory disease. We defined chemotherapy refractoriness as no achievement of at least a PR with the most recent chemotherapy containing salvage regimen before enrolling into anti-CD19 CAR-T protocol. Two of the ALL patients had extramedullary involvement.

**Table-4.** Patient Characteristics

| Patient | Disease | Pre-CAR-T Treatments | Pre-CAR-T<br>LDH/Ferritin/CRP | HSCT Before CAR-T | Bridging<br>Therapy | CAR-T Dose<br>(Total) | Response | Toxicity |
| --- | --- | --- | --- | --- | --- | --- | --- | --- |
| P1 | DLBCL<br>Bulky | RT, CHOP, R-Hyper CVAD,<br>Ibrutinib | 177/1536/8.9 | Allogenic (DLI+)<br>Post-HSCT Time: 19<br>months | RT | $5 \times 10^6$/kg | PR | CRES Grade<br>1 |
| P2 | ALL | ALL-IC BFM 2009, RT FLAG-IDA | 207/2376/3.3 | Allogenic<br>Post-HSCT Time: 14<br>months | No | $6.4 \times 10^6$/kg | CR | CRS Grade 2 |
| P3 | ALL | Hyper CVAD | 138/1541/0.19 | Allogenic<br>Post-HSCT Time: 8<br>months | Vincristine<br>Methylprednis<br>olone | $4.5 \times 10^6$/kg | PD | No |
| P4 | DHL | R-Bendamustine, ESHAP,<br>GEMOX | 13/596/1.1 | Autologus | RT | $4.8 \times 10^6$/kg | PD | No |
| P5 | DLBC<br>Bulky | R-EPOCH-R, R-ICE, RT,<br>Nivolumab, GDP | 86/2453/5.7 | Autologus | No | $4 \times 10^6$/kg | PR | CRS Grade 1 |
| P6 | DLBCL<br>Bulky | R-CHOP, ICE | 596/600/16.8 | No | No | $7.3 \times 10^6$/kg | PR | No |
| P7 | ALL | Hyper CVAD, FLAG-IDA | 260/1555/2.6 | Allogenic<br>Post-HSCT Time: 7<br>months | No | $4 \times 10^6$/kg | CR | CRS Grade 2<br>CRES Grade<br>1 |

Clinical data illustrates that the study group is very heavily pre-treated (median line of the previous therapy=4) with all except one having a previous history of HSCT, including three patients who had allogeneic HSCT. Time from HSCT to ISIKOK-19 infusion were 19,14, 8, 7 months, and donor chimerisms at the time of leukapheresis were 100, 98, 73, and 0 consecutively. One of the patients received checkpoint inhibitor therapy (Nivolumab) 6 months before ISIKOK-19 infusion. No graft-versus-host disease was observed following infusion of ISIKOK-19.

Infusion of ISIKOK-19 was associated with mild and transient toxicity; one patient had grade 1 CRS and another one had grade 1 CRES which were resolved without intervention. One patient who had grade 2 CRS, needed a single dose of tocilizumab and recovered afterward. One patient had Grade 2 CRS, Grade 1 CRES, and hematological toxicity with pancytopenia. He needed both tocilizumab and dexamethasone treatment for recovery. Hematological toxicity was resolved by the end of the second month. All four patients who had CRS or CRES were responsive to ISIKOK-19 therapy. All of the patients except one experienced B cell aplasia following ISIKOK-19 infusion. B cell aplasia lasted longer in responding patients.

All of the patients with DLBCL obtained PRs after infusion of CAR-T cells. All three of these DLBCL patients had the bulky disease at the time of CAR-T infusion. The only DHL patient had progressive disease. Two of the patients with ALL obtained CR and the other ALL patient had no response.

Patient No. 1 was diagnosed with CLL transformed to DLBCL. He underwent treatment with radiotherapy, rituximab, cyclophosphamide, doxorubicin, vincristine, and prednisone (R-CHOP); but there was progressive lymphoma. He achieved complete remission with rituximab, cyclophosphamide, doxorubicin, vincristine, dexamethasone, methotrexate, and cytarabine (R-Hyper CVAD) and underwent allogeneic HSCT from a matched sibling donor. He had a relapse after one year post-HSCT and received CHOP and ibrutinib supported by donor lymphocyte infusions. He had no response to salvage therapy. Although he received radiotherapy as bridging to CAR-T, he had a bulky cervical mass at the time of CAR-T infusion. He received 5×10^6^ cells/kg. He had somnolence on day 6 following CAR-T cell infusion and slept over 20 hours. This toxicity was evaluated as Grade 1 CRES and. recovered spontaneously without intervention. He had a partial response in the first month of infusion. (Supplementary Figure-1). However, there was a progressive disease during the third month of infusion. Biopsy of the progressive lesion showed scarce CD19 expression, unlike the initial lesion.

Patient No. 2 was diagnosed to have ALL. She received ALLIC BFM 2009 protocol and stayed in remission for one year following the end of chemotherapy protocol. She achieved remission and underwent allogeneic HSCT from a matched unrelated donor. She had a second relapse, including extramedullary and bone marrow, one year post-HSCT. She had an extramedullary mass at the right breast, multiple bone involvements, and bone marrow involvement at the time of CAR-T infusion. She received 6.4×10^6^ cells/kg. On day 4 she had refractory fever and hypotension. Single-dose tocilizumab was used on day 4 and she recovered afterward. This event was evaluated as grade 2 CRS. She had complete remission at the 4th month of ISIKOK-19 infusion and as consolidation therapy, she received a second allogeneic HSCT from another matched related donor. She had relapsed disease in the 6th month of the second HSCT and died as a result of progressive disease. (Figure-2).

Patient No. 3 was diagnosed to have ALL. She received four courses Hyper CVAD and underwent allogeneic HSCT from a matched unrelated donor. She had isolated bone marrow relapse during the sixth month of HSCT. She had no response to salvage chemotherapy and had 50% blasts at the time of CAR-T infusion. She received bridging treatment with methylprednisolone and vincristine before lymphodepletion. CAR-T infusion timing was 8 months following the allo-HSCT. Her CAR-T cells could not proliferate properly (copies per microgram of genomic DNA that show proliferation of CAR-T cells following infusion were low) and had progressive disease on the 21st day of CAR-T infusion. Although she received another salvage chemotherapy followed by a second allo-HSCT, she died due to progressive disease.

Patient No.4 was diagnosed to have follicular lymphoma. She underwent treatment with five cycles of rituximab and bendamustine. However, there was progressive lymphoma and rebiopsy was confirmed as double-hit lymphoma. She received salvage therapy with etoposide, methylprednisolone, cytarabine, and cisplatin (ESHAP) and underwent autologous HSCT. Only three months later she had relapsed disease and could not achieve remission with three cycles of gemcitabine/ oxaliplatin. She had progressive disease at the time of CAR-T infusion and had progressive disease again after CAR-T cell therapy. Following CAR-T cell therapy, partial response was achieved with methotrexate and cytarabine. She underwent allogeneic HSCT from a matched unrelated donor but died due to progressive disease at the sixth month of allogeneic HSCT.

Patient No.5 was diagnosed to have DLBCL and underwent treatment with rituximab, etoposide, cyclophosphamide, doxorubicin, vincristine, and prednisolone (R-EPOCH); the result was progressive lymphoma. He achieved remission with three cycles of rituximab, ifosfamide, carboplatin, and etoposide (R-ICE) and underwent autologous HSCT. In the third month of transplantation, he had progressive disease and received nivolumab. In the second month of nivolumab therapy, he had pericarditis (probably due to nivolumab) and progressive disease. He was treated with gemcitabine, cisplatin, and dexamethasone but was unresponsive. He had a right-sided bulky mass in his chest wall, and radiotherapy was applied as bridge treatment to CAR-T infusion. There were 6 months between the last nivolumab and CAR-T cell infusion. He received 4×10^6^ cells/kg and he had a fever on day 5 which was responsive to antipyretic drugs. This event was evaluated as grade 1 CRS and recovered spontaneously. He had a partial response in the second month of CAR-T therapy. (Supplementary Figure 3). Three months following CAR-T cell treatment, he was hospitalized because of fever, pancytopenia, and respiratory failure. New lesions and pleural effusion were detected and after a diagnostic work-up (including biopsy, cultures, imaging, and PCR), lymphoma, COVID-19, and other infections were excluded. Galactomannan was high in bronchoalveolar lavage fluid and he showed a response to anti-fungal treatment. In the 6th month of CAR-T therapy, he presented with fever and respiratory failure again. Biopsy was reperformed from the mediastinal lesion and this time it was consistent with NHL. Salvage treatment was started for the relapsed disease but the patient died due to progressive disease.

Patient No.6 was diagnosed to have DLBCL and underwent treatment with three cycles of R-CHOP and two cycles of ICE; the result was progressive lymphoma. He was referred for CAR-T therapy with a bulky mass at the left inguinal region. Early PET/CT analysis at day 16 was done because of the increasing size of the bulky mass and it was consistent with progression. Conversely, at the routine control PET/CT on day 28, there was a partial response in comparison with lesions at the beginning of CAR-T therapy (Supplementary Figure-4). Regarding the response on day 28, the results on day 16 were accepted as pseudo-progression. The patient still had a partial response in the second month of CAR-T cell therapy but died due to COVID-19 pneumonia.

Patient No. 7 was diagnosed to have ALL. He received Hyper CVAD chemotherapy, but later had isolated bone marrow relapse and received salvage chemotherapy. He achieved remission and underwent haploidentical HSCT from his parent. He had combined bone marrow and CNS relapse at the 6th month of HSCT. He was administered CAR-T cells at the dose of 4×10^6^ cells/kg. He was pyrexial during the first 48 hours, followed by liver function impairment and somnolence. With a diagnosis of Grade 2 CRS and Grade 1 CRES and he was administered tocilizumab on day 5. Dexamethasone was added to treatment on day 7 because of refractory fever and pancytopenia. The patient had complete remission in the third month of treatment. In summary, 9 patients were enrolled for the trial (ALL n=5 and NHL n=4) but only 7 patients could receive the treatment. (Table-5). Two out of three ALL patients and three out of four NHL patients had complete/partial response (ORR 72%). Four patients (57%) had CAR-T-related toxicities (CRS, CRES, and pancytopenia). Two patients were unresponsive and had progressive disease following CAR-T therapy. Two patients with partial response had progressive disease during follow-up. One patient with partial response died due to COVID-19 infection. One of the two patients with complete response underwent hematopoietic stem cell transplantation although she had detectable CAR-T cells and B cell aplasia at the time of transplantation. Unfortunately, she relapsed following the transplantation. The other patient with complete remission is still on follow-up with good performance status.

**Table-5.** Clinical Outcomes According to Diagnosis

| Diagnosis | Number of Patients | Number of Treated Patients | Number of Bridging Therapy | ORR | CR | PR | NR | Toxicity |
| --- | --- | --- | --- | --- | --- | --- | --- | --- |
| ALL | 5 | 3 | 1 | 67% | 2 (67%) | - | 1 (33%) | 2 CRS (67%),<br>1 CRES (33%) |
| NHL | 4 | 4 | 2 | 75% | - | 3 (75%) | 1 (25%) | 1 CRS (25%),<br>1 CRES (25%) |

## Discussion

CAR-T cell manufacturing has many potential pitfalls that could occur at any step from leukapheresis to the final product leading to failure of the administration of the final product ^12-14^ Results of this study are therefore promising from a productional point of view. In this study, eight aphereses for eight patients were performed and only one production failed and one patient died just before CAR-T cell infusion. Seven of the eight attempts resulted in a successful outcome that met all the criteria for releasing and quality control. The product had another advantage in terms of cost-effectiveness, as previously demonstrated by Ran et al. who reported low cost of decentralized CAR T-cell production in an academic nonprofit setting.^15^ Hence, in comparison with commercialized CAR-T cells, academic CAR-T cell academic production is less costly and hence more available to the population. Despite reduced expansion capacity of this group of heavily pretreated patient’s T-cells, satisfactory product potency in terms of cytotoxic potential and pro-inflammatory cytokine production was observed.

The ratio of certain T cell subsets in the final CAR-T product may influence the efficacy of treatment. It was shown that T_N_, T_SCM,_ and T_CM_ phenotypes were associated with higher anti-tumor efficacy and longer persistence.^11, 16, 17^ In this study, T_N_, T_SCM,_ and T_CM_ phenotypes of one of the two non-responders were below the mean value of the entire group. In this study, there was a high degree of T cell variability and CD45RO positive memory phenotype was dominant in the final product. Also, regarding CD4+ and CD8+ phenotypes, a greater CD4+ expansion was observed in comparison with CD8+ expansion ex vivo, with the exception of one patient who had also T cell proliferation problems. This is in agreement with previous reports.^6, 18^

This study presents the data of a heavily pre-treated, end-stage group of patients who had active and/or bulky disease at the time of CAR-T cell infusion. Despite the challenging disease status, there was a 72% response rate with an acceptable toxicity profile.

Although the response rate in our study was comparable to similar studies^19-21^, the median duration of response was unsatisfactory with only 5 months. Progressive disease was observed 4 and 5 months following treatment in two patients who initially had a partial response. On the other hand, one of the responsive patients died very early (after 2 months) due to COVID-19 infection. The patient who had the longest duration of response was the ALL patient with complete response to CAR-T cell therapy, who could be bridged to HSCT, but she relapsed afterward. It is controversial to perform HSCT after CAR-T cell therapy and the complete response of this patient may have persisted without HSCT as she had detectable CAR-T cell and B cell aplasia at the time of HSCT.

Lasting high CAR-T copy numbers and vanishing CD19 positivity in responsive patients were evidence for the efficacy of ISIKOK-19 cells. CAR-T copies disappeared only following consolidation HSCT in the patient with complete response, and CAR-T copies were still high for the responsive lymphoma patients at the time of progression. In the responsive lymphoma patients, biopsy specimens after disease progression showed no CD19 positive cells, suggesting that progression occurred in the CD19 negative lymphoma cell population in given patient. These findings were consistent with the high rates of CD19 negativity in relapses and these relapses mostly occurred within the first 4 months following CAR-T cell therapy.^22^

Poor prognostic factors were defined in CAR-T cell therapy previously. High tumor burden is one of the prominent factors associated with poor prognosis.^23^ The mechanism could be explained by challenging the tumor microenvironment and also by immundysregulation induced by a high tumor burden.^24^ Although it is known that response rates are not satisfactory in DLBCL, response rates in this study (3 PR for 3 patients) for bulky DLBCL appears acceptable.^25, 26^

Efficacy of CAR-T therapy in extramedullary disease in ALL is unclear as yet, but preliminary studies reported promising results.^27, 28^ In this study, one of the ALL cases who had breast and bone involvement at presentation, had a complete response and was successfully bridged to HSCT at the fourth month of CAR-T therapy. The other ALL patient with CNS involvement was still in remission at the third month of the CAR-T treatment. This is an encouraging finding for CAR-T cell use in extramedullary ALL relapses.

Four out of seven patients in this study had a history of allogeneic HSCT before CAR-T cell infusion. Three of these four patients had a high ratio of donor chimerism (100%, 98%, and 73%) at the time of leukapheresis and one of them was non-chimeric. The patients with high donor chimerism showed response while the non-chimeric patient was unresponsive to treatment. This is in accordance with previous CAR-T reports from patients with high donor chimerism.^29^ We speculate that satisfactory results with high donor chimeric cases may have been due to the non-exhausted T cells of donors in comparison with the probably exhausted T cells of heavily pre-treated patients. This data is supportive for allogenic CAR-T cell trials and encouraging with mild toxicity and no graft versus host disease.

Checkpoint inhibitor experience in CAR-T cell therapy is another current matter of debate. Potential induction of alloimmunity is the most feared complication of checkpoint inhibitors and the safe timing of use is not yet defined. One such patient in this study, where the last dose of nivolumab was used approximately six months prior to CAR-T cell infusion, no complication was observed and the patient had a partial response to CAR-T cell treatment. A recent study reported that the use of checkpoint inhibitors just before CAR-T cell infusion was safe and efficient.^30^

Bridging therapies were used for three patients in this study (chemotherapy for one patient and radiotherapy for 2 patients) before CAR-T cell infusions. There is a wide spectrum of bridging therapy options in the literature.^31^ Radiotherapy as a bridging treatment in CAR T-cell therapy is being researched^32^ and one such patient with DLBL had a partial response and no adverse events due to radiotherapy were observed in the two patients in this study.

Overall, the toxicity observed in the study was mild. Grade 2 CRS was noted in two patients who required intervention. All responders had adverse events whereas non-responders had no toxicity. This finding supports the observation about the correlation between adverse events and the efficacy of CAR-T cell therapy in previous reports.^33, 34^ It is also known that a high tumor burden is associated with more adverse events in CAR-T cell therapy.^35^ Low toxicity rates in this study which mainly included patients with high tumor burden can be explained by split dose application over three days. All patients with the exception of one (who also had the lowest CAR-T cell expansion) experienced B cell aplasia at various time points following CAR-T cell infusion. This data supports the efficacy of ISIKOK-19 as there has been a close correlation between B cell aplasia and the anti-tumoral capacity of CAR-T cells.^33^

The limitation of this study was the inclusion of mainly heavily pre-treated refractory patients. This influenced the clinical outcome in this small group of patients. Furthermore, COVID-19 pandemic period led to the death of one of the study patients. The findings of this study need to be supported by the current ongoing larger clinical trial of ISIKOK-19.

## Conclusion

This study presented the productional and clinical outcomes of the first CAR-T cell trial in Turkey. Production efficacy and fulfilling the criteria of quality control were satisfactory for academic production. Response rates and toxicity profiles are acceptable for this heavily pretreated/refractory patient group. ISIKOK-19 cells appear to be a safe, economical, and efficient treatment option for CD19 positive tumors.

## Data Availability

All data are available via Acibadem Hospital's electronic medical records system.

## Acknowledgement/Disclaimers/Conflict of interest

None

