## Supplementary Figure 1 for "Preliminary Report of Academic CAR-T (ISIKOK-19) Cell Clinical Trial in Turkey: Characterization of Product and Outcomes of Clinical Application"

Supplementary Figure 1: Cervical CT images of Patient 1 before CAR-T cell therapy (up) and at day 28 of CAR-T cell therapy (below)

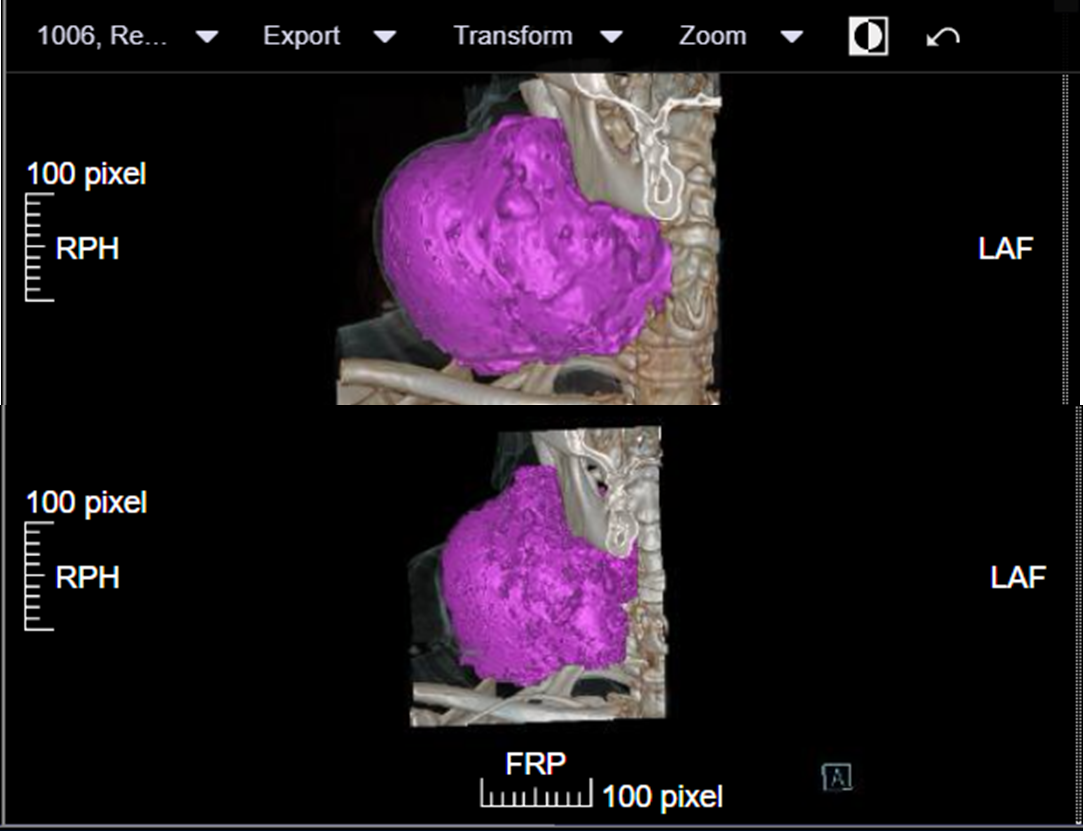
