## Supplementary Figure 3 for "Preliminary Report of Academic CAR-T (ISIKOK-19) Cell Clinical Trial in Turkey: Characterization of Product and Outcomes of Clinical Application"

Supplementary Figure 3: PET/CT of Patient 5 before CAR-T cell therapy (left) and at the second month of CAR-T cell therapy (right)

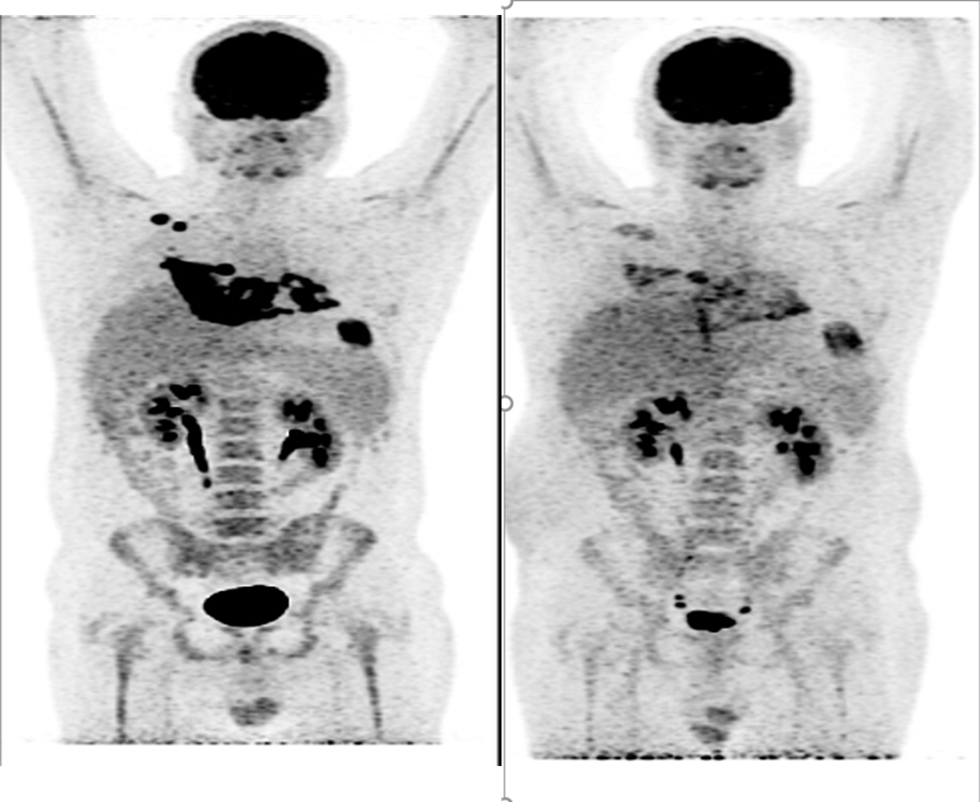
