## Supplementary Figure 4 for "Preliminary Report of Academic CAR-T (ISIKOK-19) Cell Clinical Trial in Turkey: Characterization of Product and Outcomes of Clinical Application"

Supplementary Figure 4: PET/CT of Patient 6 before CAR-T cell therapy (left) and at day 28 of CAR-T cell therapy (right)

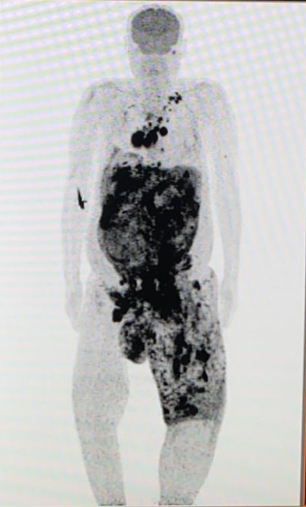

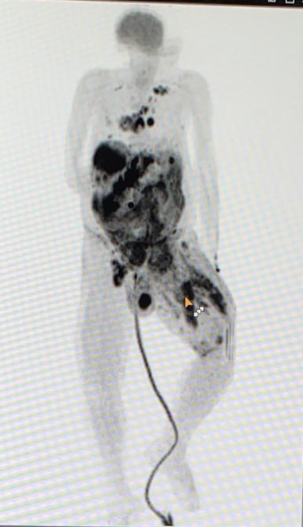
